# Socioeconomic and Clinical Determinants Driving Access to BRCA Genetic Testing in Cancer : A Case-Control Study Using Observational Electronic Health Records Across Multiple Sites

**DOI:** 10.64898/2026.05.14.26353261

**Authors:** Qiang Yang, Chengrong Wang, Charite Ricker, Sandra G. Suther, Qianqian Song, Sarah Khan, Yi Guo, Thomas J George, Mattia Prosperi, Rui Yin

**Affiliations:** Department of Health Outcomes and Biomedical Informatics, University of Florida, College of Medicine, Gainesville, FL 32610, USA; Keck School of Medicine, University of Southern California, Los Angeles, CA 90033, USA; College of Pharmacy and Pharmaceutical Sciences, Institute of Public Health, Florida Agricultural and Mechanical University, Tallahassee, FL 32307, USA; UF Health Breast Center, University of Florida, Jacksonville, FL 32209, USA; Department of Medicine, Division of Hematology & Oncology, University of Florida, Gainesville, FL 32610, USA; Department of Epidemiology, College of Public Health and Health Professions, College of Medicine, University of Florida, Gainesville, FL 32610, USA

## Abstract

**Importance:** *BRCA* genetic testing is critical for cancer risk assessment, treatment and personalization, yet substantial underutilization persists. Socioeconomic and clinical factors may strongly influence testing uptake; therefore, identifying the potential drivers to *BRCA* testing and treatment is essential for addressing gaps in access, increasing retention into care, and improving cancer outcomes.

**Objective:** To quantify the putatively causal effects of SDoH on *BRCA* genetic testing among individuals with breast, ovarian, pancreatic, and prostate cancers and to develop a predictive model to identify patients at risk for underuse of testing.

**Design, Setting, and Participants:** This observational case-control study used data from a large multistate clinical research data network covering southern US (2012-2023). The network contained records of more than 26 million individuals and was linked with ZIP code-level SDoH variables derived from national socioeconomic datasets. Adults diagnosed with breast, ovarian, pancreatic, or prostate cancer were eligible for cases (received BRCA testing) or controls (did not receive BRCA testing, matched by cancer diagnosis).

**Exposure:** SDoH categories, including economic conditions, education, healthcare access, neighborhood conditions, and social connectedness.

**Main Outcomes and Measures:** The primary outcome was receipt of *BRCA* genetic testing after cancer diagnosis.

**Results:** Among 3,279 people diagnosed with cancer, 748 received *BRCA* testing and 2,531 served as controls. Study population’s mean [SD] age was 66.8 [15.7] years; 1,758 were women [53.6%], 2,238 [69.6%] were White and 616 [18.8%] were Black or African American. Breast (1,420 [42.8%]) and prostate (1,342 [40.9%]) cancers were the most common diagnoses, followed by pancreatic (242 [7.4%]), ovarian (238 [7.2%]), and multiple cancers (55 [1.7%]). Upon adjusting for potential confounding, higher educational attainment (odds ratio [OR], 1.19), public-sector employment (OR, 1.42), neighborhood safety (OR, 1.28), and social participation (OR, 1.72) showed an increased likelihood of undergoing *BRCA* testing, whereas economic instability, including housing cost burden and reliance on public insurance, had an effect of reduced testing. A random forest classifier demonstrated good discriminative performance (AUROC, 0.776) to predict cancer patients who were likely to take *BRCA* testing, where nativity, language, and residential stability ranked among the most influential social determinants according to SHapley Additive exPlanations (SHAP) analysis.

**Conclusions and Relevance:** In this observational case-control study, SDoHs were strongly associated with receipt of *BRCA* genetic testing among people with cancer. These findings suggest that disparities in genetic testing may reflect structural and social barriers rather than differences in clinical eligibility alone. Efforts to improve equitable access to genetic testing may benefit from integrating social-context information into clinical workflows and targeting outreach or navigation strategies toward socially disadvantaged populations.

**Key Points:** *Question:* Do socioeconomic and clinical circumstances contribute to care access gaps to *BRCA* genetic testing among people with breast, ovarian, prostate, and pancreatic cancers?

*Findings:* In a case-control study of 3,279 adults with cancer from a large multi-state US clinical research data network, multiple social determinants of health, including economic conditions, education, healthcare access, neighborhood conditions, and social connectedness, were identified as key determinants of *BRCA* genetic testing uptake, with variation across age and sex groups.

*Meaning:* Improving equal access to *BRCA* genetic testing may require interventions that address financial barriers and strengthen social, and community supports that influence engagement with genetic services.

## Introduction

Genetic testing plays a crucial role in identifying individuals at an increased risk of developing certain types of cancer, particularly for *BRCA1* and *BRCA2* mutations^1–3^. Pathogenic variants in *BRCA1/2* confer significantly increased mortality risks for breast, ovarian, pancreatic, and prostate cancers, and their early identification enables timely risk-reducing interventions and tailored therapeutic strategies^4,5^.

Despite clear clinical utility and National Comprehensive Cancer Network (NCCN) guidelines recommending universal or risk-based *BRCA* testing^6–9^, the utilization in real-world practice remains low and unevenly distributed^10,11^. Studies have shown that 50–80% of eligible people do not undergo *BRCA* testing^12^. This is especially pronounced among individuals with lower socioeconomic status, rural populations, and in generally underserved demographic groups^13–17^. Mounting evidence suggests that social determinants of health (SDoH), including income, education, healthcare, employment, and neighborhood context, play critical roles in shaping access to genetic testing^18–20^. Investigating the underlying causal factors is essential to reduce health gaps, ensure equal access to genetic testing, and strengthen cancer care pathways from early detection through treatment.

In this study, we aim to systematically examine the potential socioeconomic and clinical factors that lead to disparities in *BRCA* testing for four cancer types (i.e., breast, ovarian, pancreas, and prostate) using large-scale patient-centered profiles from a large (>26 million people) multi-state clinical data research network, covering Southern USA. We propose to combine causal inference and machine learning^21^ to identify and evaluate how these factors influence *BRCA* testing utilization in cancer patients. Our findings would not only generate more precise insights into the root causes of healthcare gaps but also inform targeted interventions and policy changes to improve access across heterogeneous subpopulations.

## Methods

### Data Source and Study Design

We conducted an observational case-control study using data from the OneFlorida+ consortium, part of the US National Patient-Centered Clinical Research Network (https://pcornet.org/). The study was approved by the University of Florida Institutional Review Board (NO. IRB202400752). OneFlorida+ covers over 26 million patients across Florida, Alabama, Georgia, Arkansas, California, and Minnesota, collating their deidentified electronic health records (EHRs), comprehensive of individual-level demographics, diagnoses (encoded via the International Classification of Diseases, ICD), procedures (Current Procedural Terminology, CPT), medications (RxNorm), lab tests (Logical Observation Identifiers, Names, and Codes, LOINC), and residential information at the 5/9-digit ZIP code-level. SDoH variables were obtained from the American Community Survey (ACS) Data Profiles, which summarize key social, economic, housing, and demographic indicators for defined geographic areas. Data from 2012–2023 at the ZIP Code Tabulation Area (ZCTA) level were retrieved from *data.census.gov*, maintained by the U.S. Census Bureau. Area-level SDoH measures were linked to electronic health record data using a ZIP code-to-ZCTA crosswalk provided by UDS Mapper^22^. Individual-level exposure during the study period was estimated based on longitudinal residential history, using a duration-weighted averaging method^23^.

Eligible participants for our study were adults (≥18 years) diagnosed with breast, ovarian, pancreatic, or prostate cancers (ICD 10: C50.X, C56.X, C25.X, C61) between 2012 and 2023. To identify those cancer patients who received *BRCA* genetic testing, we used a comprehensive, manually curated combination of CPT and LOINC codes, provided in the **Supp. Section 2**. Codes corresponding to family-history screening and somatic or tumor sequencing were excluded, as this study focused specifically on germline *BRCA* testing performed in the context of a personal cancer diagnosis for hereditary risk assessment and clinical management. Patients tested prior to their cancer diagnosis were additionally excluded to ensure the outcome reflected testing initiated as part of cancer care rather than pre-diagnostic hereditary screening. Controls were selected from patients with cancer who did not receive *BRCA* testing and were matched to cases in a 5:1 ratio based on year of cancer diagnosis. The workflow of study sample construction is shown in **Figure 1A**.

**Figure 1:**
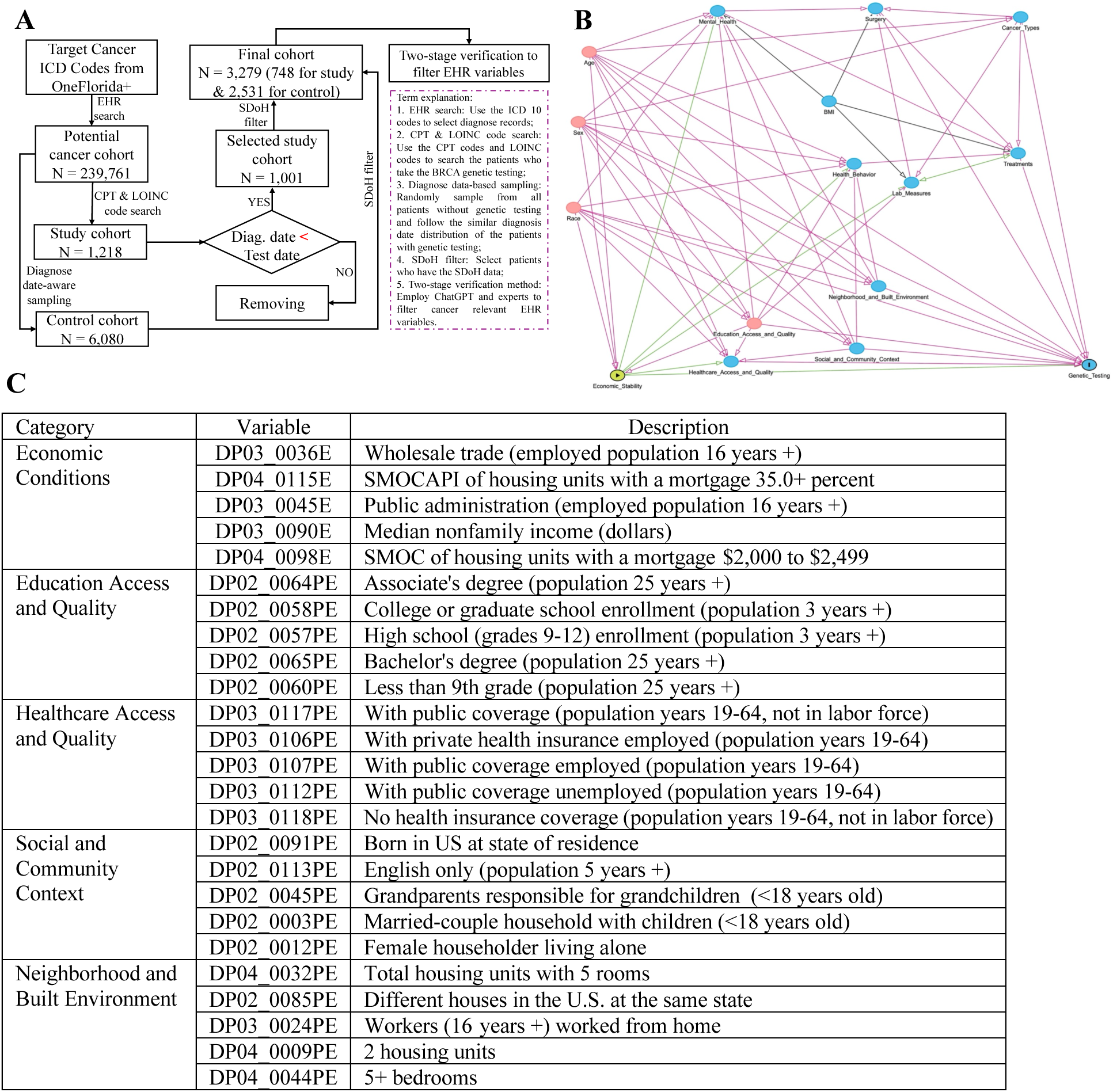
Overview of data preparation and analytical workflow. (**A**) Cohort construction process for identifying eligible cancer patients and defining case-control groups. (**B**) Example directed acyclic graph (DAG) illustrating economic conditions as the exposure and the corresponding adjustment set, including age, sex, race, and education access and quality. (**C**) Final set of selected social determinants of health (SDoH) features included in the analysis.

### Study Variables and Data Processing

We extracted demographic (e.g., age, sex, and race), clinical (e.g., laboratory results, mental health indicators, alcohol use, vaccination status, vital signs, and medication history), and SDoH variables from the electronic health records. SDoH variables were grouped into five categories, economic conditions, education access and quality, health care access and quality, neighborhood and built environment, and social and community context, according to the Healthy People 2030 framework^1^. For brevity, these categories are referred to as economy, education, healthcare, neighborhood, and social.

To mitigate redundancy and instability arising from highly correlated socioeconomic variables, we implemented a structured variable selection procedure at both the category and individual-variable levels. Initially, variables with more than 20% missing values were excluded to reduce instability associated with extensive imputation. For the remaining variables, simple imputation was applied prior to correlation-based grouping to preserve the observed correlation structure among SDoH variables used for clustering and variable selection. Continuous variables were imputed using the median or mean depending on variable type, while binary variables were imputed using the mode (majority class). Multiple imputation was not performed because the primary objective of this step was to maintain the empirical correlation structure among SDoH variables used for variable grouping. Within each SDoH category, pairwise correlations were calculated, and hierarchical clustering was performed on the absolute correlation matrix using average linkage. For clusters of highly correlated variables ((|*r*| > 0.8), the variable with the strongest absolute association with the outcome was retained as the cluster representative. If the number of retained variables exceeded a predefined threshold (e.g., 5 variables per category), further reduction was performed by selecting those with the largest absolute correlations with the outcome. This procedure resulted in a final set of 20 individual-level SDoH variables, which were combined with adjustment covariates for downstream modeling (**Figure 1C**).

### Causal Inference Modeling

We employed directed acyclic graphs (DAGs) and the generalized adjustment criterion (GAC)^24^ to estimate the total effects of category-level and individual-level SDoH variables on the receipt of *BRCA* genetic testing. DAGs were constructed based on domain knowledge and expert review to specify assumed causal relationships among exposures, outcome, and covariates. For each SDoH, the GAC identified a minimal sufficient adjustment sets to block backdoor paths while avoiding collider or overadjustment bias. For example, a DAG used to analyze economic conditions identified age, gender, race, and educational opportunities and quality as confounding factors that need to be adjusted for (**Figure 1B**). DAGs and corresponding adjustment sets for all other SDoHs are provided in **Supplementary Figure 1** and **Supplementary Table 2**.

Causal effects were estimated using generalized linear models (GLM)^25^ with a logit link function, with receipt of *BRCA* genetic testing as a binary outcome. Each individual-level SDoH variable was standardized (z-scored) prior to model fitting to enable comparability of effect sizes across variables. Binary SDoH variables were retained in their original 0/1 form to preserve interpretability of the corresponding model coefficients. For category-level analyses, composite scores were constructed for each SDoH category by calculating the mean of standardized individual variables within that category. Each GLM included a single SDoH variable as the exposure and its corresponding DAG-derived minimal sufficient adjustment set calculated via the GAC. Effect estimates were reported as adjusted odds ratios (ORs) with 95% confidence intervals.

To assess heterogeneity across demographic groups, analyses were stratified by age group (<40, 40–59, 60–79, and ≥80 years) and sex group (female and male), and subgroup-specific odds ratios were estimated. To formally evaluate effect modification, interaction terms between each SDoH variable and the stratification variable were included in separate multivariable models, and the statistical significance of the interaction term was used to test whether associations differed across groups on the multiplicative scale. All analyses were performed using R version 4.4 (*R* Foundation for *Statistical Computing, Vienna, Austria*). GLMs were fitted using the *stats* package. Data manipulation was performed using the *tidyverse* suite of packages.

### Machine Learning Prediction

While the causal analysis quantifies the average effects of SDoHs on *BRCA* genetic testing receipt, with some information about heterogeneity through stratification, it does not provide individualized risk prediction. To complement these analyses, we developed a machine learning model to predict the likelihood of receiving *BRCA* genetic testing and to identify the most influential predictors through model interpretation, although these predictors may not necessarily have a causal meaning, for which we have performed the other analysis.

Prior to model training, a feature preprocessing pipeline was implemented to remove low-information and redundant features among socioeconomic and clinical variables. A two-stage feature filtering strategy was applied. It the first stage, domain knowledge–driven selection was used to identify feature categories likely to be relevant to the cancers of interest and *BRCA* genetic testing, informed by literature and expert input (e.g., educational attainment in SDoH variables and cancer-related medications in clinical variables). In the second stage, data-driven statistical filtering was performed. Variance thresholding was applied to remove near-zero variance features. Pearson correlation filtering was used to retain features with sufficient association with the outcome. Univariate statistical tests were then conducted to assess feature relevance, including ANOVA F-tests for continuous variables and chi-square tests for binary variables. Features not meeting predefined criteria were excluded. In total, 313 variables were retained for model development.

A random forest classifier^26^ was developed to predict the likelihood of receiving *BRCA* genetic testing using the prefiltered demographic, clinical, and SDoH features. The dataset was randomly partitioned into training (80%) and testing (20%) sets. Predictive performance was evaluated on the held-out test set using area under the receiver operating characteristic (AUROC), sensitivity, specificity, and F1 score. To enhance interpretability, SHAP^27^ values were computed to quantify the contribution of each input feature to the predicted probability *of BRCA* genetic testing. SHAP values estimate the impact of individual features on model predictions by calculating their average marginal contribution across all possible feature combinations, thereby identifying the most influential variables driving model predictions. SHAP summary plots were generated to visualize global feature importance and the directionality of associations with *BRCA* testing utilization. Machine learning analyses were performed using Python 3.10 with *scikit-learn* for preprocessing and model training, and the SHAP library for model interpretation.

## Results

### Characteristics of the study sample

The final study population included 3,279 patients, of whom 748 received *BRCA* testing (cases) and 2,531 did not (controls). As shown in **Figure 2A**, age distribution was similar between the two groups, with the majority of people aged 60-70 years, indicating an older population typical of cancer people. Gender composition differed modestly between groups, with comparable proportions of women (1,333 vs 425) and men (1,198 vs 323) in the control and study groups (**Figure 2B**). The racial distribution was similar across cohorts (**Figure 2C**), with White participants compromising the majority (1,764 vs 519), followed by Black or African American (444 vs 172). Cancer type distributions differed between groups (**Figure 2D**). Breast and prostate cancers were the most prevalent malignancies in both cohorts, accounting for 283 and 245 cases in the study cohort (*n* = 748) and 1,119 and 1,097 cases in the control cohort (*n* = 2,531), respectively.

**Figure 2:**
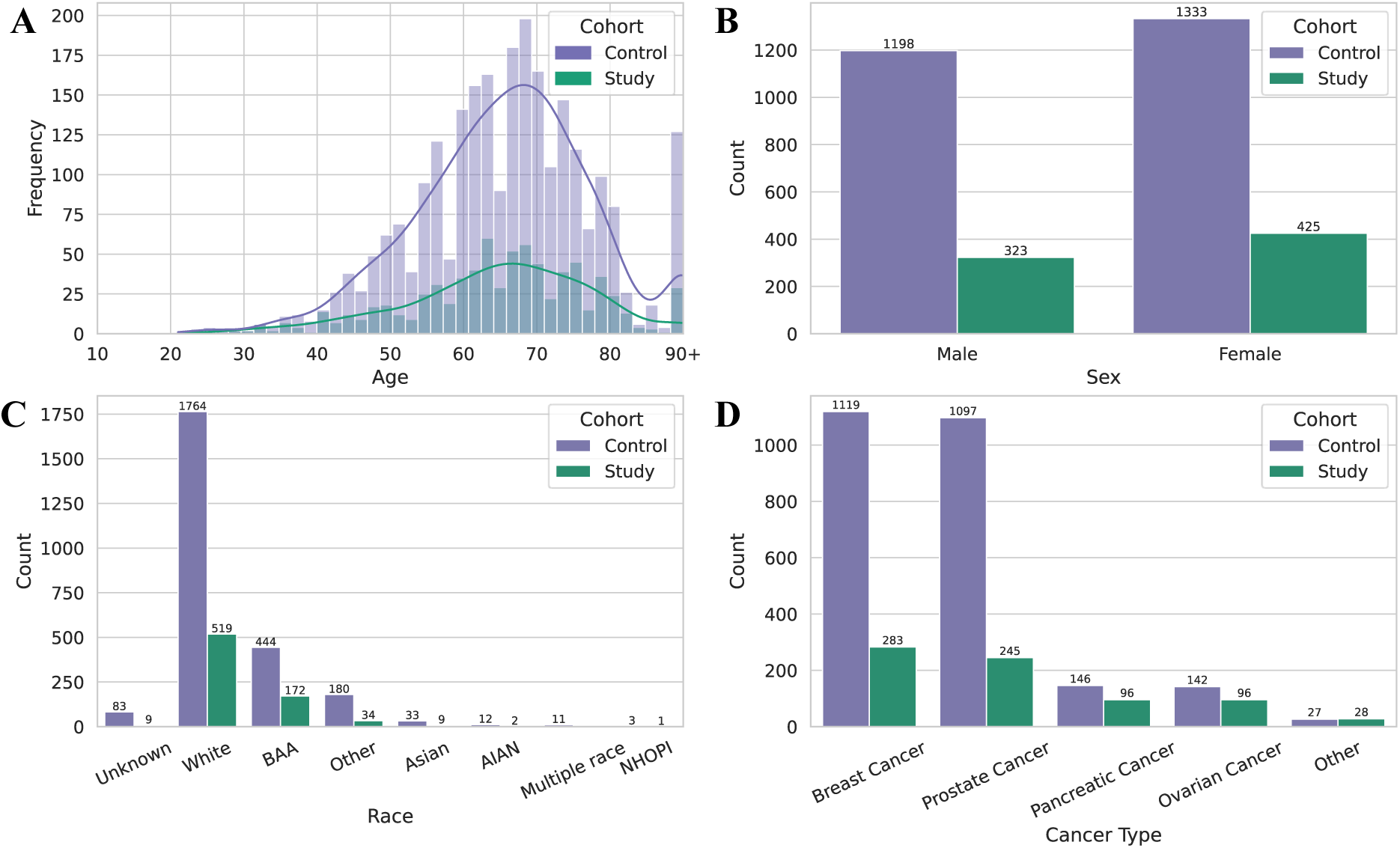
Distributions of demographic and clinical characteristics for the study and control cohorts across (**A**) age; (**B**) sex; (**C**) race; (**D**) cancer type.

### Effects of SDoHs on BRCA Genetic Testing Utilization

DAG-informed generalized linear models (GLMs) were used to estimate putative causal effects of SDoH on *BRCA* genetic testing (**Figure 3**). Accordingly, **Figure 3A** presented individual-level causal estimates of SDoH on *BRCA* genetic testing with adjusted odds ratios. Within the economic conditions category, markers of financial strain were associated with substantially lower *BRCA* genetic testing odds, including lower “median nonfamily income” (DP03_0090E; OR, 0.61; 95% CI, 0.55–0.67; p-value, 3.84e-21), higher “housing cost burden” (DP04_0098E; OR, 0.66; 95% CI, 0.59–0.74; p-value, 1.85e-13; DP04_0115E; OR, 0.67; 95% CI, 0.61–0.74; p-value, 2.54e-17), and “employment in wholesale trade” (DP03_0036E; OR, 0.59; 95% CI, 0.53–0.65; p-value, 8.60e-26). In contrast, “employment in public administration” (DP03_0045E) was associated with increased *BRCA* genetic testing (OR, 1.34; 95% CI, 1.24–1.45; p-value, 8.12e-14). In the education access and quality, “college or graduate enrollment” (DP02_0058PE; OR, 1.19; 95% CI, 1.10–1.29; p-value, 6.20e-06) was positively associated with *BRCA* testing, whereas lower educational attainment, including “less than ninth grade” (DP02_0060PE; OR, 0.86; 95% CI, 0.78–0.94; p-value, 1.03e-03) and “associate’s degree prevalence” (DP02_0064PE; OR, 0.81; 95% CI, 0.74–0.88; p-value, 5.15e-07) exhibited reduced testing rate. Additionally, we found healthcare access and quality showed modest effects. Specifically, “public insurance coverage” among employed or unemployed adults (DP03_0107PE; OR, 1.12; DP03_0112PE; OR, 1.12; p-value, 4.86e-02) increased testing probability, while lack of insurance (DP03_0118PE, OR, 0.90; 95% CI, 0.82–0.99; p-value, 3.30e-02) showed a decrease for the testing. Within the neighborhood and built environment, residential stability variables, including “same-state residence” (DP02_0085PE; OR, 1.21; 95% CI, 1.11–1.31; p-value, 5.64e-06) and larger housing units (DP04_0032PE; OR, 1.19; 95% CI, 1.09–1.30; p-value, 2.14e-04), were associated with higher testing rate, whereas “remote work prevalence” (DP03_0024PE; OR, 0.85; 95% CI, 0.76-0.95; p-value, 3.44e-03) and constrained housing size (DP04_0044PE; OR, 0.89; 95% CI, 0.80-0.99; p-value, 3.90e-02) were associated with lower chance. When we looked at the social and community context, the results exhibited the strongest positive effects on *BRCA* testing. We observed that “English-only households” (DP02_0113PE; OR, 1.82; 95% CI, 1.63–2.04; p-value, 4.67e-25), “being born in the state of residence” (DP02_0091PE; OR, 1.72; 95% CI, 1.57–1.89; p-value, 1.23e-31), and “grandparent caregiving” (DP02_0045PE; OR, 1.43; 95% CI, 1.31–1.55; p-value, 6.17e-17) were robustly associated with increased testing, whereas “married-couple households with children” (DP02_0003PE) were associated with lower testing odds (OR, 0.79; 95% CI, 0.72–0.86; p-value, 1.16e-07).

**Figure 3.**
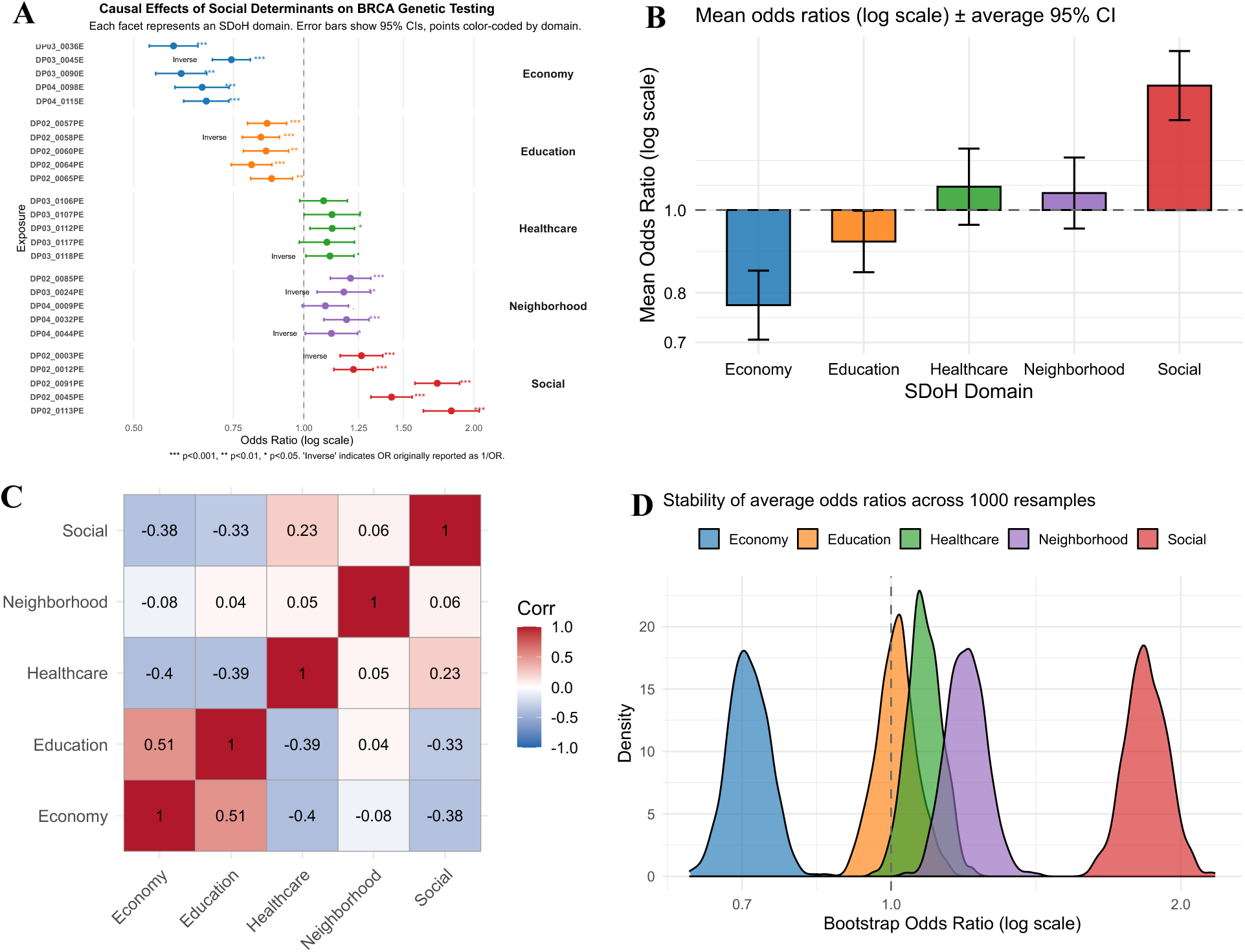
Causal analysis of social determinants of health (SDoH) and *BRCA* genetic testing. (**A**) Adjusted associations of individual-level SDoH variables with *BRCA* testing, estimated using generalized linear models (GLMs). Each line represents an odds ratio (OR) with a 95% confidence interval, color-coded by category. (**B**) Category-level summary effects, showing the overall direction and magnitude of associations. (**C**) Correlation heatmap of SDoH category scores, illustrating interrelationships among socioeconomic factors. (**D**) Bootstrap distributions (1,000 resamples) for category-level effect estimates, demonstrating robustness. Dashed vertical lines denote null effects (OR = 1), and asterisks indicate statistical significance (p < 0.05, p < 0.01, p < 0.001). Full names of SDoH variables shown in Figure 3A are provided in Figure 1C.

We additionally summarized causal effects at the category level by averaging adjusted odds ratios across individual-level SDoH variables within each category for *BRCA* genetic testing (**Figure 3B**). The economic conditions showed the lowest mean odds ratio (mean OR, 0.78), indicating that economic factors related to financial strain were associated with lower likelihood of receiving *BRCA* genetic testing. In contrast, the social and community context category exhibited the strongest positive association (mean OR, 1.18), followed by modest positive effects for the neighborhood (mean OR, 1.04) and health care access (mean OR, 1.05). The correlation matrix of the five SDoH categories was shown in **Figure 3C**. Economic conditions and education showed a moderate positive correlation (*r* = 0.51). Both domains were moderately negatively correlated with the healthcare access domain (*r* = −0.40 and *r* = −0.39, respectively). Correlations among the remaining domains were generally weak, including small positive correlations between healthcare access and social context (*r* = 0.23) and between neighborhood environment and social context (*r* = 0.06). **Figure 3D** displayed bootstrap distributions of category-level odds ratios for *BRCA* genetic testing based on 1,000 resamples. Economic conditions showed a stable negative association, whereas social and community context showed the strongest positive association. Healthcare access and neighborhood context were modestly positive, while education was centered near the null, suggesting no consistent overall association.

### Age-Stratified Analysis

We found substantial differences in how SDoH influenced *BRCA* genetic testing across age groups (**Figure 4A-C**). Economic conditions demonstrated the most pronounced age-specific variation in association with *BRCA* genetic testing **(Figure 4A)**. Among adults aged 40-59 years, variables indicative of economic disadvantage were consistently associated with substantially lower odds of *BRCA* testing (OR < 1, p < .001), indicating that financial strain represents a key barrier during working-age adulthood. In contrast, among adults aged 60-79 years, higher economic conditions were associated with increased testing uptake (OR > 1, p < .001), suggesting that economic resources facilitate access to genetic testing at older ages. Associations were weaker and less consistent among individuals younger than 40 years and those aged 80 years or older. This pattern may partly reflect differences in genetic testing guidelines across cancer types, as age historically influenced eligibility for *BRCA* testing in breast cancer but plays a smaller role for ovarian and pancreatic cancers. Across other SDoH categories, social and community context and neighborhood conditions showed modest effects in younger age groups but became increasingly positive among adults aged 60 years or older, while healthcare access exhibited a similar strengthening pattern with age for *BRCA* testing. Education showed relatively stable and modest associations for *BRCA* testing across all age groups.

**Figure 4.**
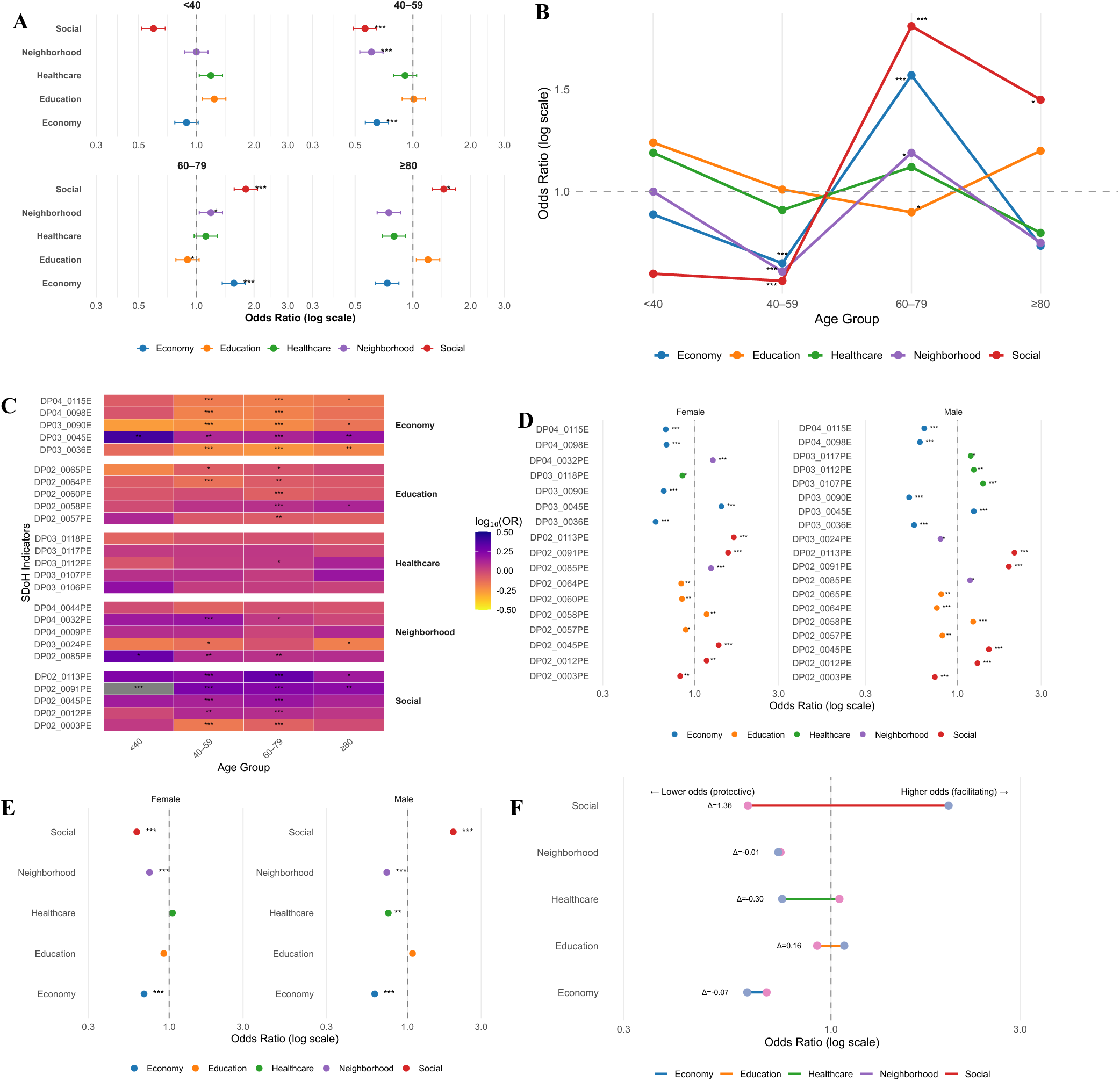
Stratified associations of SDoH with *BRCA* genetic testing by age and sex. (**A**) Category-level associations of SDoH with *BRCA* testing across age strata, estimated using GLMs. (**B**) Age related trajectories of category-level associations across the lifespan. (**C**) Heatmap of individual-level associations of SDoH with *BRCA* testing stratified by age group. (**D**) Individual-level associations stratified by sex, presented as odds ratios (ORs) with 95% confidence intervals (CIs). (**E**) Category-level associations stratified by sex, illustrating differences in effect magnitude and direction. (**F**) Dumbbell plot comparing male versus female adjusted ORs across SDoH categories, highlighting sex-specific differences in associations. Full names of individual-level SDoH variables shown in **panels C** and **D** are provided in Figure 1C.

**Figure 4B** depicted age-specific causal effects of SDoH on BRCA genetic testing at category level, where we observed that economic conditions emerged as the strongest barrier among adults aged 40-59 years (mean OR, 0.60), with this negative association attenuating in older adults and partially reversing among those aged 60-79 years (mean OR, 1.30) for *BRCA* testing. In contrast, social and community context became the dominant facilitator with increasing age, rising from modest effects in midlife to strong positive associations for *BRCA* testing in adults aged 60-79 years (mean OR, 1.70) and those aged 80 years or older. Neighborhood conditions and healthcare access demonstrated smaller but progressively positive effects on *BRCA* testing after age 60 (ORs, 1.10-1.30), while education showed relatively stable and modest associations across age groups.

**Figure 4C** presented a heatmap illustrating age-stratified causal effects of individual-level variables on *BRCA* genetic testing. We found that among adults aged 40-59 years, several variables reflecting economic burden were strongly associated with lower likelihood of *BRCA* testing, including wholesale trade employment (DP03_0036E; OR, 0.60; p < .001), lower median nonfamily income (DP03_0090E; OR, 0.57; p < .001), and housing cost burden (DP04_0115E; OR, 0.67; p < .001). In adults aged 60-79 years, public-sector employment (DP03_0045E; OR, 1.32; p < .001) and college or graduate school enrollment (DP02_0058PE; OR, 1.19; p < .001) were associated with higher testing rates. The neighborhood stability, including “residential mobility within the same state” (DP02_0085PE) and “housing size” (DP04_0032PE), showed consistent positive associations with *BRCA* testing across age groups. Additionally, social context has demonstrated the strongest positive effects on the *BRCA* testing, particularly “US nativity” (DP02_0091PE), “English-only language use” (DP02_0113PE), and “grandparent caregiving” (DP02_0045PE), with odds ratios exceeding 1.3 among adults aged 40-79 years. In conclusion, economic status most affects the chances of *BRCA* testing in midlife groups, whereas social integration and neighborhood stability can promote the testing rate in elder groups.

### Sex-Stratified Analysis

We found distinct patterns in how socioeconomic conditions influenced *BRCA* genetic testing (**Figure 4D-F**). At the individual variable level (**Figure 4D**), economic instability was consistently associated with lower odds of *BRCA* testing in both women and men. Variables of economic disadvantage, including lower-wage employment (e.g., “wholesale trade employment” [DP03_0036E: OR, 0.60 in females; OR, 0.57 in males]) and housing cost burden (e.g., “high mortgage cost burden” [DP04_0115E: OR, 0.69 in females; OR, 0.65 in males]) were associated with reduced odds of *BRCA* testing. “Public administration employment” (DP03_0045E) was positively associated with *BRCA* testing in both sexes, with a stronger effect observed among females (OR, 1.42) than males (OR, 1.24). Educational variables showed modest associations in both groups, while healthcare access measures, such as “public insurance coverage” among the employed (DP03_0107PE: OR, 1.40) and “unemployed individuals” (DP03_0112PE: OR, 1.24), were more consistently associated with *BRCA* testing among males.

At the category level (**Figure 4E**), association between SDoH categories and *BRCA* genetic testing were directionally consistent across sexes but differed in magnitude. Economic instability remained a strong barrier to *BRCA* testing in both males (OR, 0.62; p < .001) and females (OR, 0.69; p < .001). The social and community context showed the strongest positive association overall, with a larger effect in males (OR, 1.98; p < .001) than females (OR, 1.62; p < .001). Healthcare access exhibited a sex-specific pattern, with a negative association among males (OR, 0.75; p, .003) but no significant association among females (OR, 1.05; p, 0.54). Education and neighborhood categories showed smaller and comparable effects across sexes. The dumbbell plot (**Figure 4F**) highlighted sex-based differences by directly comparing adjusted odds ratios across SDoH categories. The largest difference occurred in the social and community context, where social cohesion and community engagement were more strongly associated with *BRCA* testing in males than females (*Δ*OR = +0.36). In contrast, healthcare access showed an inverse pattern, with lower testing odds among males relative to females (*Δ*OR = −0.30). Differences in education were modest (*Δ*OR ≈ +0.16 favoring females), while economic conditions (*Δ*OR = −0.07) and neighborhood conditions (*Δ*OR ≈ −0.01) showed minimal sex differences. In conclusion, these findings indicate that while the same SDoH categories influence *BRCA* genetic testing in both sexes, the likelihood for receiving *BRCA* genetic testing in male patients is more dependent on social-context factors, whereas testing among females is relatively more influenced by healthcare-access and educational factors.

### Individual Risk Prediction and Feature Importance

**Figure 5A** presented classification metrics for random forest models trained on different data modalities. The model using combinations produced the highest overall accuracy (0.793), indicating more balanced identification of patients who did and did not receive *BRCA* testing. Models using SDoH data alone showed the highest sensitivity (0.527), although the magnitude of this improvement was modest. In contrast, models based on laboratory data alone achieved the highest specificity (0.907), reflecting stronger identification of patients who did not receive *BRCA* testing. Similarly, the model incorporating all available data achieved the highest discriminative performance, i.e., area under the receiver operating characteristic curve (AUROC, 0.776), followed by models using laboratory data alone (AUROC, 0.714) and SDoH data alone (AUROC, 0.701). However, the demographics-only model showed limited predictive ability (AUROC, 0.493). These results indicate that social and clinical data contributed complementary information for predicting *BRCA* testing receipt.

**Figure 5.**
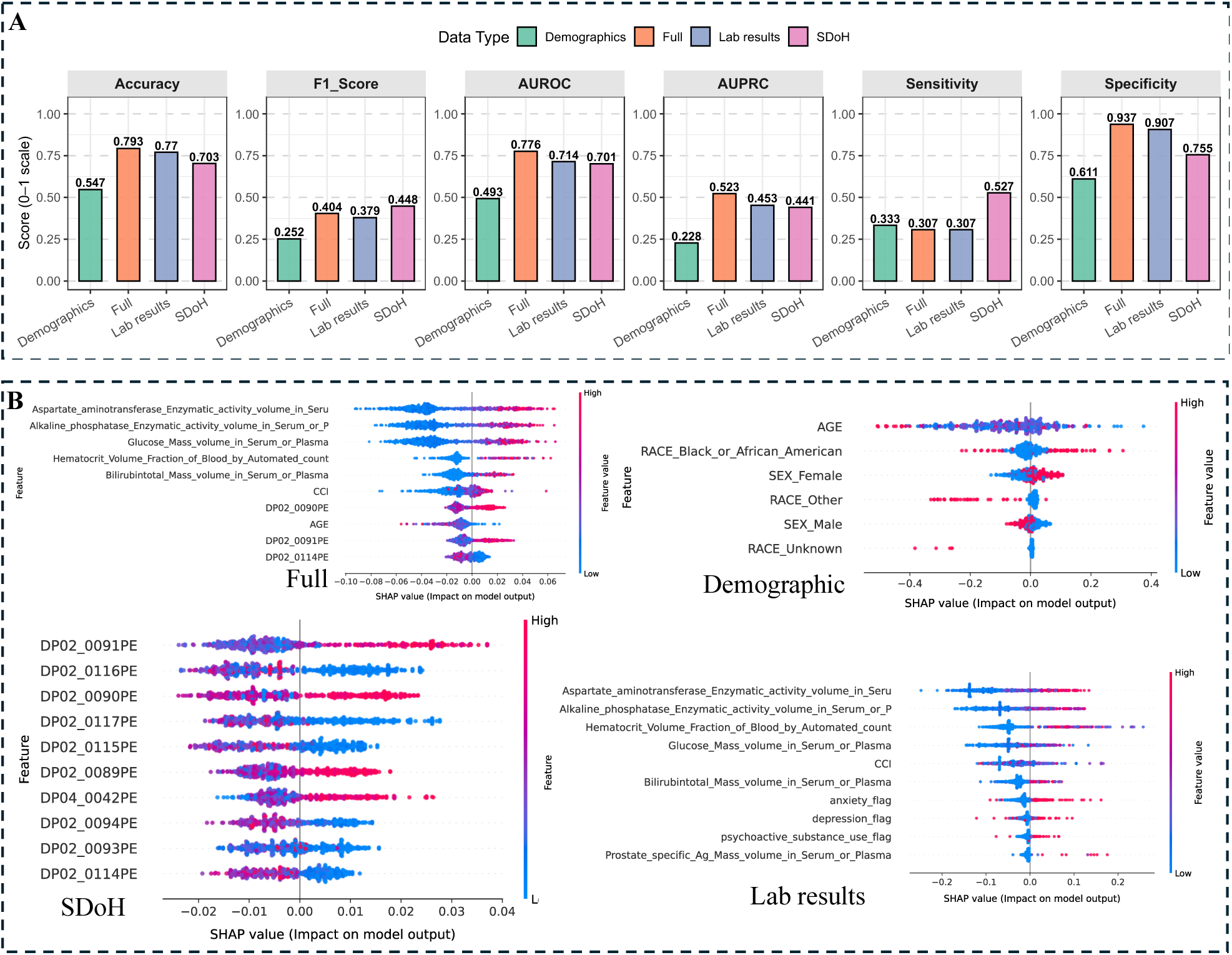
Predictive model performance and interpretability for *BRCA* genetic. (**A**) Comparison of model performance different combinations of input data. (**B**) SHAP-based feature importance illustrating the relative contribution of predictors to model outputs. Full names of SDoH variables are provided in Figure 1C.

**Figure 5B** summarized SHAP values from the random forest model across four data modalities, highlighting the relative contributions of variables to predictions of *BRCA* genetic testing utilization. In the demographic panel, age was the most influential variable, with sex and race-related variables also contributing to model predictions. The SDoH panel indicated that features related to nativity, language, and residential stability, such as “US nativity (DP02_0091PE) “, “birth in state of residence” (DP02_0090PE), and limited English proficiency (DP02_0114PE, DP02_0117PE), ranked among the most influential social determinants, with substantial variability across individuals. The model using lab results showed that biomarkers indicating liver function and metabolic or hematologic status (e.g., bilirubin, alkaline phosphatase, hematocrit, and glucose) had strong contributions to predicted testing likelihood. In the model involving all the variables, social determinants related to nativity and language emerged as the strongest predictors: higher proportions of “US-born individuals” (DP02_0091PE) and those “born in their state of residence” (DP02_0090PE) were associated with increased predicted testing likelihood, whereas greater “non-English language use at home” (DP02_0114PE) was associated with lower likelihood. Age and selected laboratory measures also contributed substantially, highlighting the combined influence of social context and clinical profiles on *BRCA* testing utilization prediction.

### Comparison of Causal Inference and Machine Learning Analyses

Results from causal inference and machine learning analyses were largely concordant, identifying overlapping SDoH as key factors associated with *BRCA* genetic testing while providing complementary perspectives. Causal analyses quantified population-level effects, showing that social and community context, particularly indicators of residential stability such as birth in the state of residence (DP02_0091PE) and intergenerational caregiving, was associated with higher testing utilization, whereas economic instability and housing burden were consistent barriers. In contrast, machine learning models emphasized predictive relevance and individual-level heterogeneity. SHAP analyses identified similar social-context variables, including residential stability, nativity, and language environment, as among the most influential contributors to predicted testing likelihood. Importantly, these findings do not imply causal inference from the machine learning models; rather, they indicate that predictive signals align with the associations observed in the causal analyses.

## Discussion

In this large case-control study, we found that several variables of financial strain, including lower median nonfamily income, high housing cost burden, and employment in lower-wage sectors, were associated with lower odds of receiving *BRCA* genetic testing. In contrast, social integration, residential stability, and public-sector employment were associated with higher *BRCA* testing. These findings suggest that differences in *BRCA* testing are influenced not only by clinical eligibility or guideline awareness but also by broader social conditions that shape access to healthcare^28–30^. By examining a very comprehensive set of individual-level and category-level SDoH, our study extends prior evidence demonstrating the underuse of *BRCA* genetic testing among socially disadvantaged populations. Correlation analysis among SDoH domains further showed that economic conditions and education were moderately positively correlated, whereas both domains were moderately negatively correlated with the healthcare access domain. These correlations describe relationships among socioeconomic domains and do not imply causal effects on *BRCA* testing utilization. Consistent with previous research, financial barriers, insurance complexity, and competing socioeconomic demands may limit engagement with genetic services even when testing is clinically indicated^6,31–33^. Conversely, employment in public administration and other variables of stable institutional access were associated with higher *BRCA* testing rates, suggesting that greater healthcare system engagement and coverage stability may partially mitigate these barriers^34^.

Social and community context was the most influential facilitator of *BRCA* genetic testing. Variables of social integration and geographic stability, particularly being born in the state of residence, were strongly associated with increased testing utilization. This pattern aligns with prior evidence that continuity of residence, language concordance, and familiarity with local healthcare systems can facilitate engagement with specialized services, including genetic counseling and testing^35–37^. It may also reflect stronger local social support networks and greater family involvement in caregiving, which could promote awareness of hereditary cancer risk and encourage participation in recommended genetic testing. Stratified analyses further revealed substantial heterogeneity in how SDoH influenced *BRCA* testing across age and sex groups. Economic barriers were most pronounced among adults aged 40 to 59 years, whereas social and neighborhood factors were more strongly associated with *BRCA* testing among adults aged 60 years or older. Sex-based analyses indicated that men were more sensitive to economic and healthcare-access constraints, while women appeared to benefit more from social cohesion and educational resources. Similar age- and sex-related differences in access to cancer-related services have been reported previously and highlight the importance of tailoring interventions to specific population subgroups rather than relying on uniform strategies^38,39^.

The concordance between causal inference and machine learning analyses strengthens the robustness of these findings. Causal models identified social context, particularly state-level nativity, as a key facilitator of *BRCA* testing, while machine learning models independently ranked the same variable among the most influential predictors at the individual level. Such alignment between causal and predictive approaches has been increasingly recognized as essential for advancing precision medicine and public health^21,40^. From a clinical and policy perspective, when an important outcome predictor is also found to be causally linked, it can be actioned through targeted programs. Interventions that reduce financial barriers, simplify insurance navigation, and strengthen community-based support may increase *BRCA* testing utilization for patients with cancer. Integrating SDoH-informed risk stratification EHR systems could further enable clinicians to identify socially disadvantaged yet clinically eligible patients and support targeted referral, counseling, or care-navigation strategies^41^.

### Limitations

This study has several limitations. First, the variables with substantial missingness were excluded, and remaining missing values were handled using simple imputation, which may introduce bias. Meanwhile, feature selection to harmonize inputs across models may omit informative variables. Second, although some SDoH variables were measured at the individual level, many were derived from the ZIP code-level data and may not fully capture individual socioeconomic circumstances. Third, residual confounding may persist despite the use of causal modeling approaches and directed acyclic graph-based confounder selection. Fourth, temporal misalignment between SDoH measurement and the timing of *BRCA* testing decisions may limit causal interpretation, as socioeconomic conditions may change over time. Fifth, changes in clinical guidelines over the study period (2012-2023) may have influenced *BRCA* testing eligibility and uptake.

## Conclusions

In this case-control study, access to *BRCA* genetic testing among individuals with cancer was shaped not only by clinical factors but also by broader socioeconomic conditions. Variables of financial strain and limited healthcare access were associated with reduced testing, whereas social integration and structural stability were linked to higher uptake. These findings suggest that underuse of *BRCA* testing is not solely a function of clinical eligibility or awareness but reflects systemic barriers embedded in patients’ social environments. Improving equitable access to genetic testing will likely require strategies beyond guideline expansion, including reducing financial and logistical barriers, strengthening care navigation and referral pathways, and leveraging community and social support systems to promote engagement with genetic services.

## Data Availability

The data can be accessed by requesting from OneFlorida+ clinical research network.

## Author Contributions

Yang and Yin had full access to all of the data in the study and takes responsibility for the integrity of the data and the accuracy of the data analysis.

Concept and design: Guo, Prosperi, Yin, Prosperi.

Acquisition, analysis, or interpretation of data: Yang, Yin.

Drafting of the manuscript: Yang, Wang, Prosperi, Yin.

Critical review of the manuscript for important intellectual content: Ricker, Suther, Song, Khan, Guo, Chu, George, Prosperi.

Statistical analysis: Yang.

Administrative, technical, or material support: Yang, Wang.

Supervision: Yin.

## Acknowledgements

Research reported in this publication was supported in part by the OneFlorida+ Clinical Research Network, 1 of 8 clinical research networks, funded by the Patient-Centered Outcomes Research Institute numbers CDRN-1501-26692, RI-CRN-2020-005, RI-FLORIDA-01-PS1 and RI-FLORIDA-01-PS8; in part by the University of Florida Clinical and Translational Science Institute, which is supported in part by the NIH National Center for Advancing Translational Sciences under award number UL1TR001427, UL1TR000064 and UM1TR005128. The content is solely the responsibility of the authors and does not necessarily represent the official views of the Patient-Centered Outcomes Research Institute (PCORI), its Board of Governors or Methodology, the OneFlorida+ Clinical Research Network, the UF-FSU Clinical and Translational Science Institute, or the National Institutes of Health.

## Supplementary Materials

### 1. Cancer Patient Characteristics in the OneFlorda+ Clinical Research Network

Using International Classification of Diseases (ICD) diagnostic codes for the four target malignancies, a total of 239,761 unique patients with cancer were identified within the OneFlorida+ clinical research network, including breast (n = 105,367), ovarian (n = 14,096), pancreatic (n = 20,763), and prostate (n = 102,431) cancers (**Supplementary Table 1**).

Age distributions reflected established cancer epidemiology. Breast and ovarian cancers were diagnosed at relatively younger ages, with 44.6% of breast cancer patients aged 45 to 64 years and 12.5% of ovarian cancer patients younger than 45 years. In contrast, pancreatic and prostate cancers were more prevalent among older adults, with 45.3% and 52.8% of patients aged 75 years or older, respectively. The proportion of patients younger than 45 years was low for pancreatic (2.9%) and prostate (0.5%) cancers.

Sex distributions aligned with the biological characteristics of each malignancy. Female patients predominated in the breast (98.3%) and ovarian (99.6%) cancer cohorts, whereas prostate cancer occurred almost exclusively in male patients (99.2%). The pancreatic cancer cohort showed a balanced sex distribution (50.9% male; 49.1% female).

Racial composition indicated that White patients were the predominant group across all cancer types (range, 61.8%–67.6%), followed by Black or African American patients (range, 15.4%–20.6%), with the highest proportion observed in prostate cancer. Asian patients accounted for 1.7%–3.4% of each group. American Indian or Alaska Native (AIAN) and Native Hawaiian or Other Pacific Islander (NHOPI) populations were minimally represented, each comprising less than 0.3% of the cohort. Patients categorized as other race or with missing racial information accounted for an additional 8.5%–11.8%.

### 2. Codes for BRCA genetic testing

To identify patients who received BRCA genetic testing, we applied a code-based algorithm informed by prior literature^42,43^ and publicly available resources from the Centers for Medicare & Medicaid Services (CMS) ^2,3,4^. First, Current Procedural Terminology (CPT) codes from the procedure table were used to identify BRCA testing, including *81162–81167, 81211–81217, 81432, 81433, 0137U, 0138U, 0172U, 83890–83892, 83896, 83903, 83912, 83914,* and *S3818–S3823*. Codes related to family-history testing or non-targeted assays were excluded to minimize misclassification. Second, Logical Observation Identifiers Names and Codes (LOINC) from the laboratory results table were used to capture BRCA-related tests, including *81247-9, 81248-7, 81249-5, 81250-3, 81251-1, 81252-9, 94231-6, 94232-4, 51966-0,* and *51967-8*. Using this approach, a total of 1,218 patients with BRCA genetic testing were identified.

### 3. Directed acyclic graphs for exposure variables

Directed acyclic graphs (DAGs) were constructed based on prior literature and expert knowledge to represent the assumed causal relationships among social determinants of health (SDoH) domains, including education access and quality, health care access and quality, neighborhood and built environment, and social and community context (**Supplementary Figure 1**). Adjustment sets derived from these DAGs are presented in **Supplementary Table 2**. Across SDoH domains, age, sex, and race were consistently identified as key confounders.

### 4. Predictive performance and interpretability using XGBoost

**Supp. Figure 2** highlighted the predictive performance and interpretability of the XGBoost across data modalities. In **Panel A**, we concluded that the full model achieved the best overall performance (accuracy = 0.796; F1 = 0.547; AUPRC = 0.534), demonstrating the advantage of integrating heterogeneous features. The SDoH-only model showed lower overall accuracy (0.674) but the highest sensitivity (0.573), indicating greater ability to identify patients who received testing. The lab-only model exhibited high specificity (0.834) with moderate sensitivity (0.540), suggesting conservative identification of non-tested individuals. The demographic-only model performed poorest across all metrics (accuracy = 0.508; F1 = 0.290), indicating limited explanatory value of demographic factors alone. **Panel B** presented SHAP-based feature importance across models. In the full model, key predictors included clinical biomarkers (e.g., bilirubin, hematocrit, glucose, alkaline phosphatase), comorbidity burden (CCI), and age. In the demographic model, female sex and White race contributed positively, whereas minority race was associated with lower predicted testing probability. The SDoH model highlighted education, housing stability, and economic indicators (e.g., DP02_0065PE). In the lab-only model, biochemical markers such as bilirubin and glucose were the most influential predictors.

---

**Supp. Table 1:** Summary statistics of cancer cases in the OneFlorida+ network, 2012–2023. BAA: Black or African American; NHOPI: Native Hawaiian or Other Pacific Islander; AIAN: American Indian or Alaska Native.

|  | **Breast**<br>*N* = 105,367 | **Ovarian**<br>*N* = 14,096 | **Pancreatic**<br>*N* = 20,763 | **Prostate**<br>*N* = 102,431 |
| --- | --- | --- | --- | --- |
| **Age** |  |  |  |  |
| <45 | 11,068 (10.5%) | 2,166 (12.5%) | 860 (2.9%) | 819 (0.5%) |
| 45-64 | 47,009 (44.6%) | 6,020 (33.7%) | 6,874 (22.3%) | 30,261 (14.2%) |
| 65-74 | 28,299 (26.9%) | 3,554 (25.3%) | 6,872 (29.4%) | 41,021 (32.5%) |
| >=75 | 18,991 (18.0%) | 2,356 (28.5%) | 6,157 (45.3%) | 30,330 (52.8%) |
| **Sex** |  |  |  |  |
| Male | 1,739 (1.7%) | 92 (0.4%) | 10,720 (50.9%) | 101,580 (99.2%) |
| Female | 103,628 (98.3%) | 14,019 (99.6%) | 10,042 (49.1%) | 839 (0.8%) |
| **Race** |  |  |  |  |
| BAA | 19,731 (18.7%) | 2,314 (15.4%) | 3,392 (16.2%) | 20,992 (20.6%) |
| White | 71,246 (67.6%) | 9,856 (63.8%) | 14,655 (64.7%) | 66,793 (61.8%) |
| Asian | 2,610 (2.5%) | 350 (3.4%) | 359 (2.6%) | 1,544 (1.7%) |
| Multiple race | 891 (0.8%) | 109 (0.6%) | 290 (1.1%) | 892 (0.8%) |
| NHOPI | 109 (0.1%) | 13 (0.1%) | 23 (0.1%) | 95 (0.0%) |
| AIAN | 228 (0.2%) | 41 (0.3%) | 44 (0.2%) | 208 (0.2%) |
| Other | 6,479 (6.2%) | 930 (6.6%) | 1,070 (5.2%) | 6,454 (6.3%) |
| Unknown | 4,073 (3.9%) | 483 (3.4%) | 930 (4.5%) | 5,453 (5.3%) |

**Supp. Figure 1:**
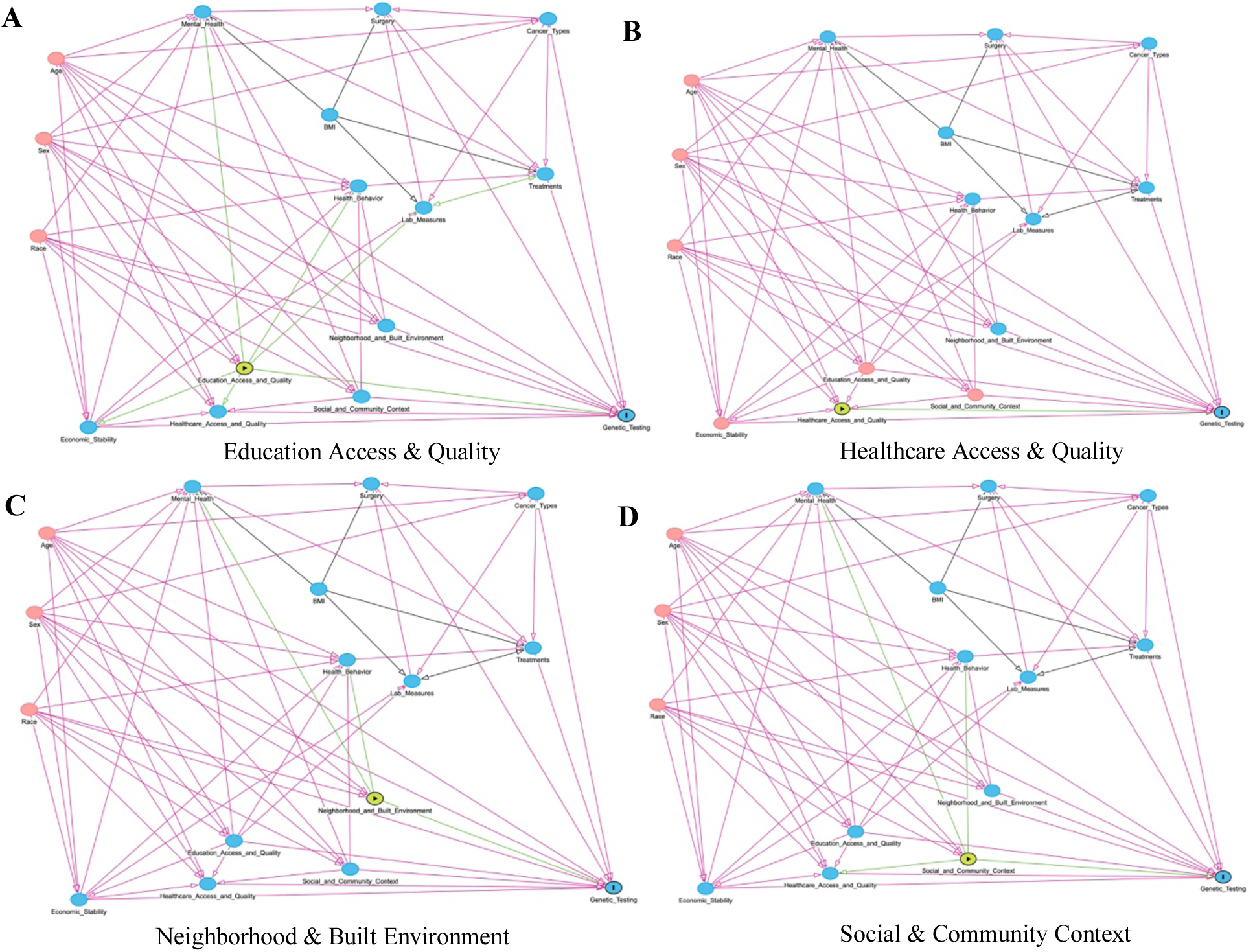
DAGs representing assumed causal relationships for each SDoH exposure category.

**Supp. Table 2:**
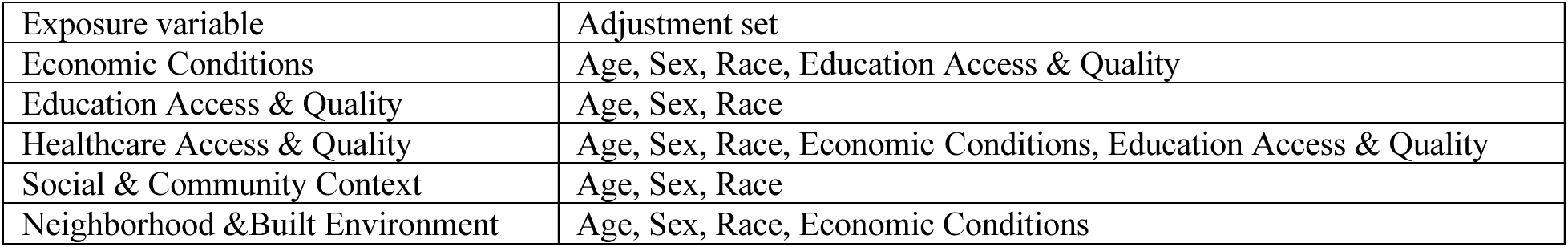
Exposure variables and corresponding adjustment sets.

| Exposure variable | Adjustment set |
| --- | --- |
| Economic Conditions | Age, Sex, Race, Education Access & Quality |
| Education Access & Quality | Age, Sex, Race |
| Healthcare Access & Quality | Age, Sex, Race, Economic Conditions, Education Access & Quality |
| Social & Community Context | Age, Sex, Race |
| Neighborhood & Built Environment | Age, Sex, Race, Economic Conditions |

**Supp. Figure 2:**
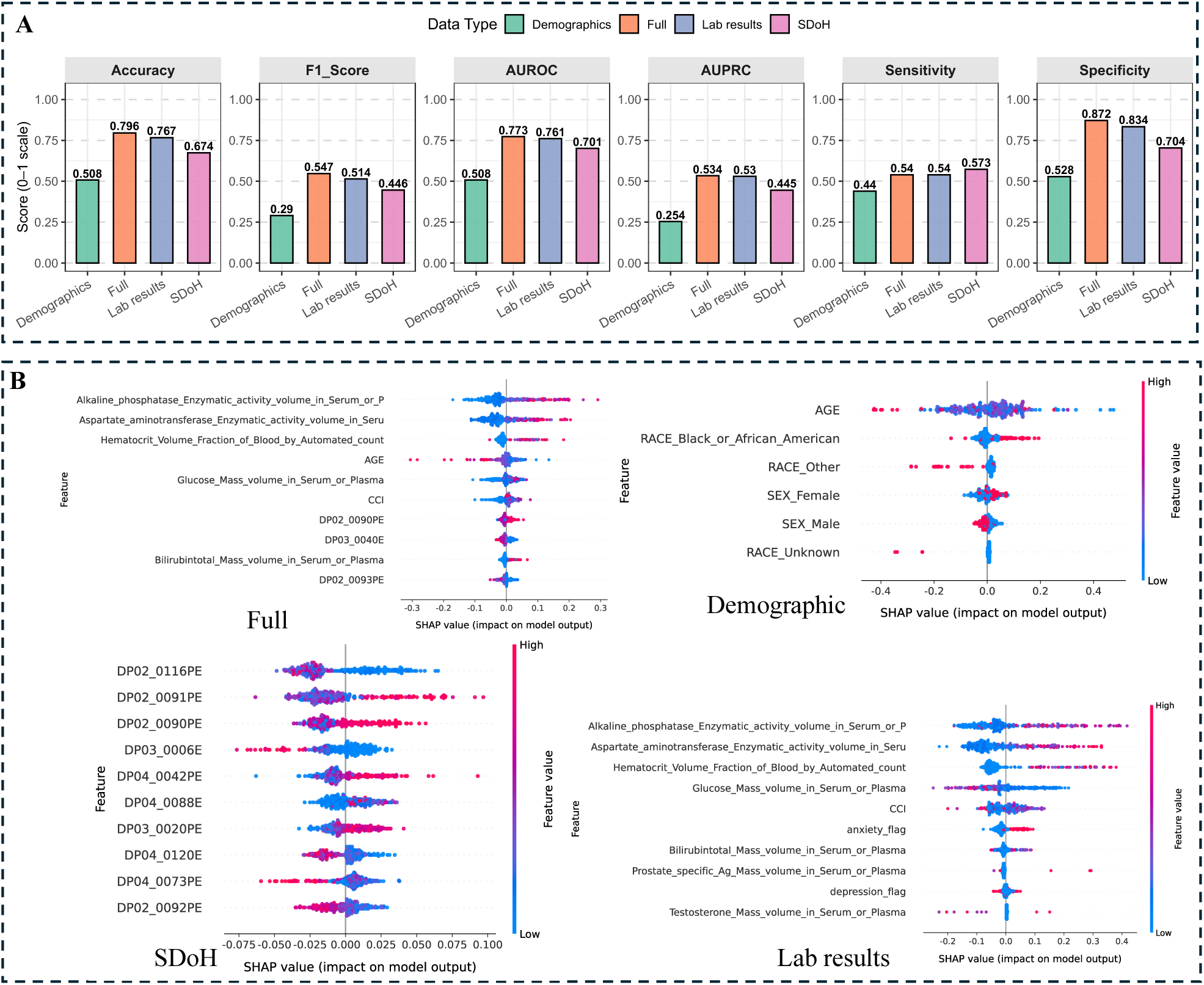
Performance and interpretability of XGBoost model across data modalities. (A) Comparison of six evaluation metrics across data types. (B) SHAP-based feature importance illustrating model interpretability. Full names of SDoH variables are provided in Figure 1C.

## Footnotes

1 https://odphp.health.gov/healthypeople/priority-areas/social-determinants-health

2 https://localcoverage.cms.gov/mcd_archive/view/lcd.aspx?lcdInfo=36714:18

3 https://www.cms.gov/medicare-coverage-database/view/article.aspx?articleId=54689&ver=32

4 https://www.cms.gov/medicare-coverage-database/view/article.aspx?articleId=56542&ver=16

